# Aging Detection Based on Dynamic State Transitions in Instantaneous Hilbert-Based Spatio-Temporal EEG Features

**DOI:** 10.1101/2025.10.22.25338412

**Authors:** Sou Nobukawa, Takashi Ikeda, Mitsuru Kikuchi, Tetsuya Takahashi

## Abstract

The spatial distribution of electroencephalography (EEG) oscillatory power and its temporal transitions are widely recognized as indicators of cognitive processes and pathological conditions, termed as microstates. These microstates reflect whole-brain neural network dynamics, including deep brain regions, and are closely associated with large-scale networks such as the default mode network. The conventional approach to microstate analysis relies on the envelope of EEG oscillations, which corresponds to the instantaneous amplitude. In this study, we aimed to extend conventional microstate analysis by integrating the instantaneous amplitude (power component) and instantaneous frequency data derived from the Hilbert transform. While our previous studies demonstrated that instantaneous frequency also reflects brain activity, this study highlights that integrating both features enables a more comprehensive assessment of aging effects. This integration allows the identification of brain states that cannot be detected using conventional power-based microstate analysis. Our findings suggest that this approach expands and enhances traditional microstate analysis and offers a novel index for detecting brain states. This method has the potential to provide new insights into neural network dynamics and can be applied to the study of cognitive processes and pathological conditions.

## 1. Introduction

Neural activity in the brain is a representative phenomenon that arises from the coordinated interaction of multiple brain regions (reviewed in [32, 2, 30, 24]). Such high-dimensional neural activity across widespread cortical and subcortical areas is not static; rather, it exhibits dynamic temporal transitions, even in the absence of external stimuli, such as during the resting state (reviewed in [7]). To capture these ongoing brain dynamics, microstate analysis has been proposed as a method of defining brain states [18, 17, 18] (reviewed in [20]). This approach focuses on quasi-stable spatial distributions of oscillatory power across brain regions and their temporal transitions. Over the past several decades, microstate analysis has been widely used to investigate the relationships between large-scale neural network dynamics, cognitive processes, and alterations associated with neurological or psychiatric conditions [36, 22, 15, 29, 38].

To capture the dynamics of large-scale brain networks, not only the power components, which have traditionally been the focus of microstate analysis, but also phase-related features (reviewed in [9, 5, 34, 35, 21]) must be considered. In particular, phase-based mechanisms have been widely utilized to describe information transfer across distributed brain regions, such as functional connectivity indices based on phase synchronization between neural activities [5] and phase-amplitude coupling, which reflect cross-frequency interactions [34]. Against this background, we previously proposed dynamic phase synchronization (DPS), a novel metric that focuses on the temporal patterns formed by phase differences between brain regions, and demonstrated its ability to detect age-related alterations in neural network dynamics with high accuracy [27]. Furthermore, we showed that the spatial distribution of the instantaneous frequency (IF) resembles that of traditional microstates derived from the envelope of neural activity, and that these IF patterns exhibit quasi-stable temporal transitions [25]. Based on these findings, we introduced the IF microstate as a new type of microstate. The dynamic properties of IF microstates are particularly relevant to pathological changes and cognitive decline in Alzheimer’s disease [26] and schizophrenia [11]. Although IF microstates exhibit spatial patterns similar to those of conventional microstates, their transition dynamics appear to be distinct, suggesting that IF microstates provide additional complementary information beyond that captured by conventional microstates [28].

During healthy aging, brain functions, such as attention, memory, and cognition, gradually decline over time [3, 23, 4, 39]. Therefore, in super-aging societies, understanding how aging alters the spatiotemporal dynamics of neural activity is critical for developing effective interventions to prevent brain dysfunction. In our previous studies, we identified age-related alterations in neural networks [13, 40, 16] by using various electroencephalography (EEG)-based approaches, including phase synchronization, temporal pattern analysis of EEG signals, and DPS [27, 1, 12, 33]. Building on these findings, we hypothesized that applying a novel method that integrates conventional microstates based on instantaneous amplitude (IA microstates) with IF microstates [26] could reveal new aspects of age-related changes in brain activity that are not detectable using either approach alone.

In this study, we propose a new microstate framework that combines the spatial distributions of both the IA and IF components derived from the Hilbert transform, which we refer to as IF-IA microstates. We then analyze the temporal transitions of these microstates during resting-state EEG in younger and older participants. Finally, we identify specific aging-related characteristics reflected in IF-IA microstates.

## 2. Materials and Methods

### 2.1 Participants

This study was conducted at the Department of Neuropsychiatry, Kanazawa University, Japan [27]. A total of 29 healthy younger participants (14 males, 15 females; mean age = 22.9 years, SD = 2.7 years; age range = 20 − 28 years) and 18 healthy older participants (7 males, 11 females; mean age = 57.5 years, SD = 4.7 years; age range = 51− 67 years) were enrolled. The groups were matched for sex (*χ*^2^ = 0.39, *p* = 0.52). All participants were nonsmokers and took no medications. Individuals with major medical or neurological conditions―including epilepsy, history of head trauma, or lifetime alcohol or drug dependence-were excluded. Additionally, participants with significant brain abnormalities (e.g., notable vascular changes observed on conventional MRI) were excluded from the study. For older participants, a Mini-Mental State Examination (MMSE) score < 27 served as an additional exclusion criterion. Written informed consent was obtained from all the participants after a full explanation of the study procedure. All methods were conducted in accordance with the Declaration of Helsinki, and the study protocol was approved by the Ethics Committee of Kanazawa University.

### 2.2 EEG Recording

EEG recordings were conducted in an electrically shielded, sound-attenuated, light-controlled room. The scalp electrodes were positioned according to the international 10–20 system. EEG data were acquired using an 18-channel system (EEG-4518, Nihon Kohden, Tokyo, Japan) from 16 electrode sites: Fp1, Fp2, F3, Fz, F4, F7, F8, C3, C4, P3, Pz, P4, T5, T6, O1, and O2, with reference to physically linked earlobe electrodes. Eye movements were recorded using bipolar electrooculography (EOG). EEG signals were sampled at 200 Hz with a time constant of 0.3 s and filtered through a 1.5− 60 Hz bandpass filter. Recordings were performed in a resting, eyes-closed condition for 10 −15 minutes. Participants were continuously monitored via video, and vigilance was assessed using EEG traces to ensure that only epochs corresponding to eyes-closed wakefulness (excluding light sleep) were analyzed. Artifacts, such as muscle activity, eye blinks, and eye movements, were visually identified. For each participant, one continuous, artifact-free 60-s epoch (12,000 data points) was extracted. Each epoch was further bandpass filtered in the theta alpha range (4 − 13 Hz) [26].

### 2.3 Microstate Analysis Based on Instantaneous Frequency and Amplitude

In this study, brain activity states were defined based on the IF and IA dynamics of the EEG signals. The estimation procedure is illustrated in Fig. 1. The multichannel EEG time series were bandpass-filtered in the range of 4 −13 Hz, which corresponds to the dominant frequency band during resting-state EEG with eyes closed. Using the Hilbert transform, the wrapped instantaneous phase *θ*(*t*) (*− π* ≤ *θ* ≤ *π*) and IA data were extracted. Because the instantaneous phase includes phase noise that may lead to large deviations in the IF estimates, known as phase slips [8], a median filter was applied to the derived IF time series. The same median-filtering process was applied to the IA signals. This estimation procedure follows the method used in our previous study on DPS [27].

**Fig. 1.**
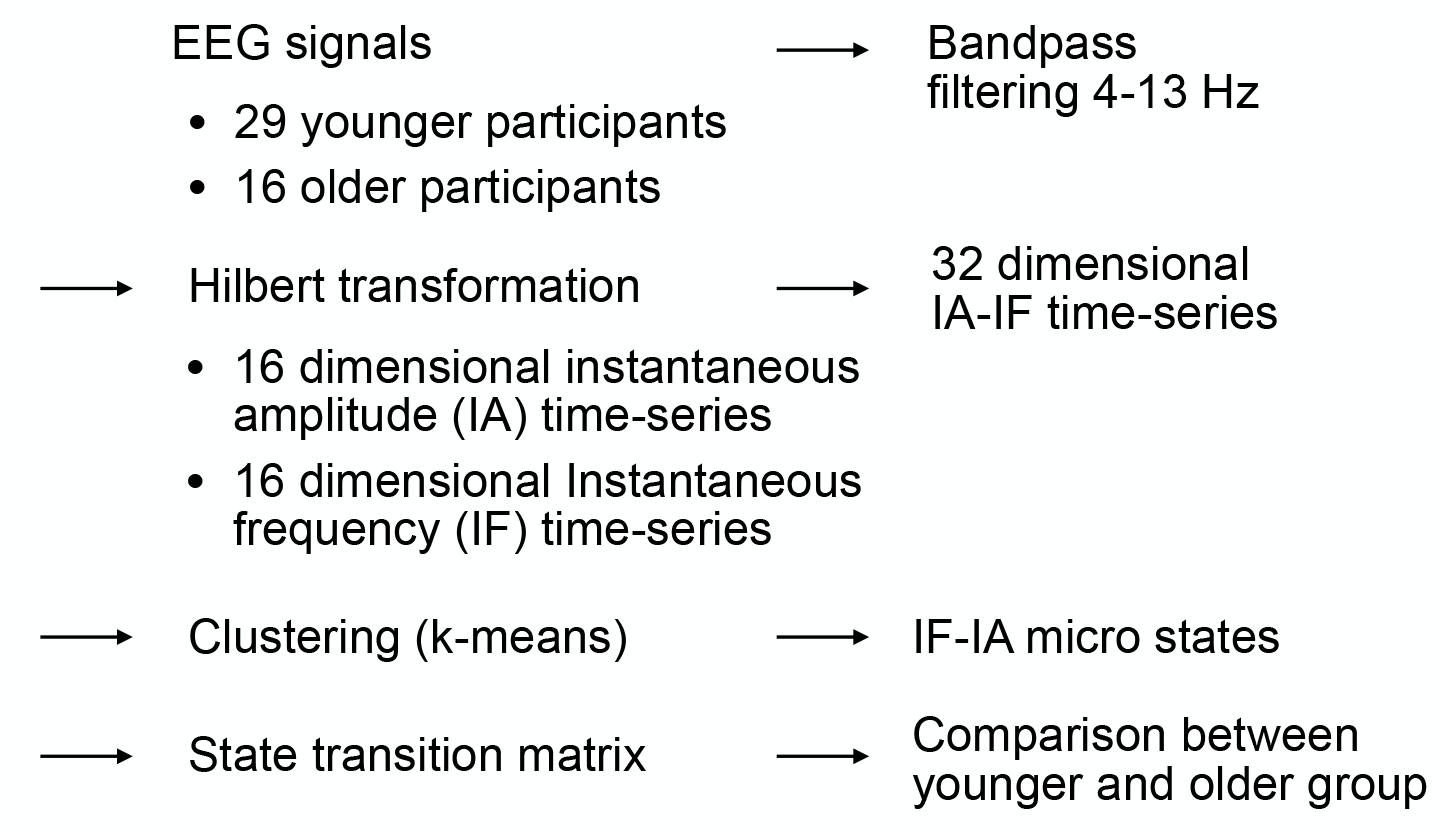
Estimation of brain states based on the instantaneous frequency (IF) and instantaneous amplitude (IA) time series derived from electroencephalography (EEG) signals.

To estimate the brain states, the spatial deviations of the IF and IA were calculated for each electrode *i*. The IF was centered by subtracting the spatial mean at each time point:

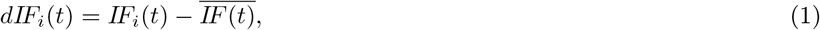

where 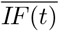 is the mean across all the electrodes at time *t*. The resulting *dIF*_*i*_(*t*) was normalized using *z*-scoring such that its standard deviation across the electrodes was 1.0. Similarly, the IA signal was centered and normalized as follows:

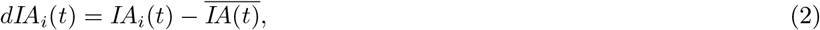

where 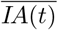 is the average IA across all the electrodes at time *t*, and *dIA*_*i*_(*t*) is also *z*-scored.

The combined time series *dIF*_*i*_(*t*) and *dIA*_*i*_(*t*) for both the younger and older groups were then clustered into *k* dynamic states using the *k*-means algorithm.Each state represents a spatial pattern composed of 32 values (16 electrodes *×* two features: IF and IA). The temporal transitions between these states were assumed to reflect moment-to-moment changes in dynamic functional connectivity (dFC) across the whole-brain network, consistent with the DPS framework. In this study, the number of clusters is set to *k* = 4.

### 2.4 Statistical Analysis

To evaluate the dynamic properties of the state transitions based on *dIF*_*i*_(*t*) and *dIA*_*i*_(*t*), we computed the transition probability matrix among the *k* states from time *t* to *t* + *Δt*, where the sampling interval is *Δt* = 0.005 s. Independent two-sample *t* tests were conducted to compare the transition probabilities between the younger and older groups. To correct for multiple comparisons across all possible state transitions (*k* × *k* = 16), the Benjamini-Hochberg false discovery rate (FDR) correction was applied (*q <* 0.05).

## 3. Results

The mean spatial deviation of the instantaneous frequency (*dIF*_*i*_(*t*)), and instantaneous amplitude (*dIA*_*i*_(*t*)) across the participants in the younger and older groups, averaged over the duration of each state, are shown in Fig. 2. In both groups, four distinct microstates were identified: occipital leading and occipital amplified (#Of-Oa), frontal leading and occipital amplified (#Ff-Oa), occipital leading and frontal amplified (#Of-Fa), and frontal leading and frontal amplified (#Ff-Fa).

**Fig. 2.**
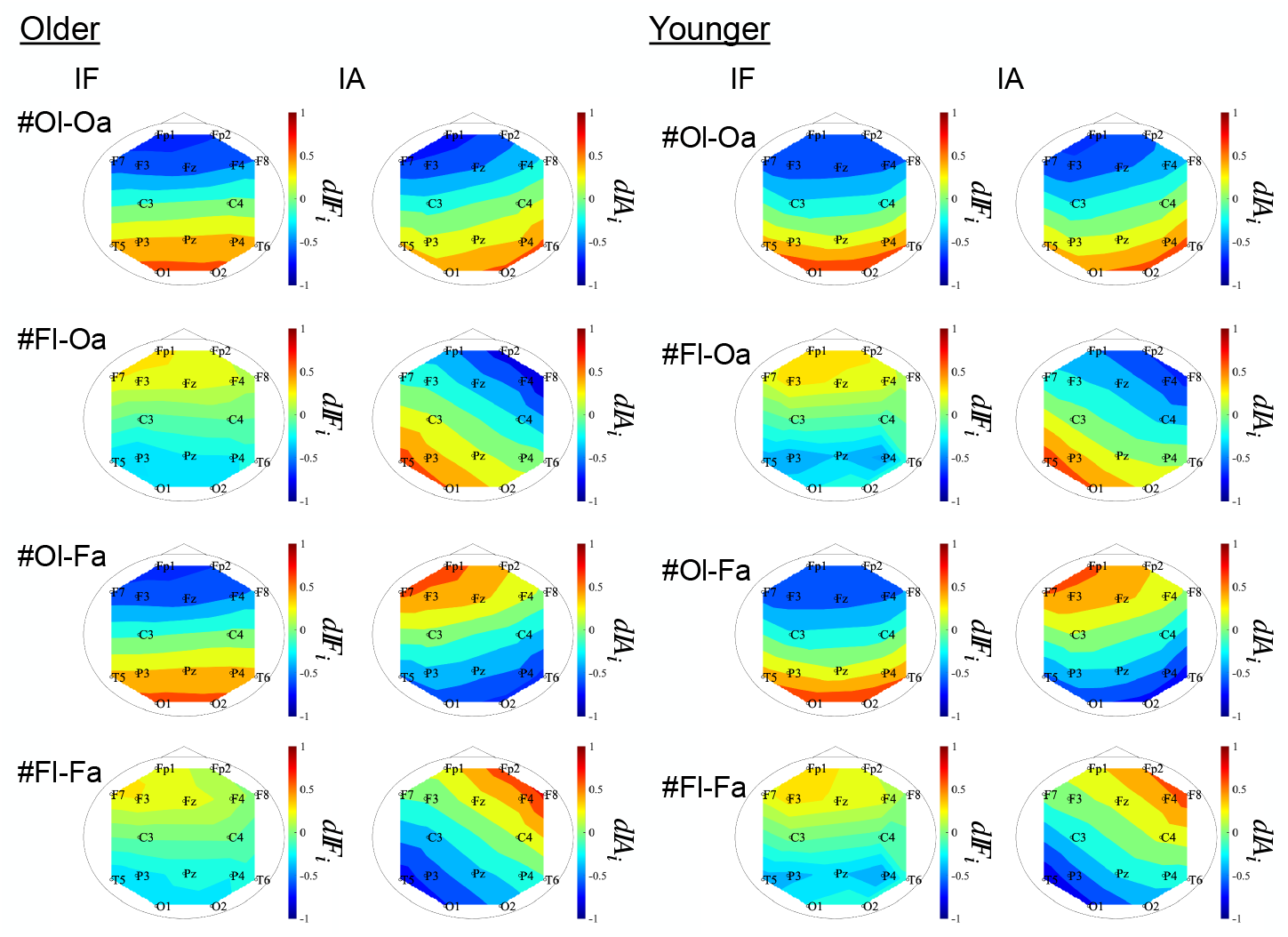
Classified spatial distributions of instantaneous frequency (IF) and amplitude (IA). The average *dIFi* and *dIAi* values within each classified state reveal four characteristic microstates: occipital-leading and occipital-amplified (#Ol-Oa), frontal-leading and occipital-amplified (#Fl-Oa), occipital-leading and frontal-amplified (#Ol-Fa), and frontal-leading and frontal-amplified (#Fl-Fa), observed in both younger and older groups.

The typical examples of time series of state transitions among #Ol-Oa, #Fl-Oa, #Ol-Fa, and #Fl-Fa are shown in Fig. 3. To capture these dynamic characteristics, Fig. 4 presents (A) the mean state transition probabilities for the younger and older groups and (B) the corresponding *t*-values of group differences. The results indicate that the transition probabilities from #Fl-Fa to #Fl-Oa and from #Fl-Oa to #Fl-Fa are significantly lower in the older group than in the younger group (*q <* 0.05). Conversely, the probability of remaining in state #Ol-Oa is significantly higher in the older group (*q <* 0.05). The probabilities of individual participants and their distributions within each group are shown in Fig. 5. Interestingly, these significant differences were detected only when *dIF*_*i*_(*t*) and *dIA*_*i*_(*t*) were analyzed simultaneously. When either component was analyzed alone (that is, *dIF*_*i*_(*t*) or only *dIA*_*i*_(*t*)), these differences were not statistically significant. This finding suggests that integrating both features is essential to accurately capture age-related alterations in state dynamics.

**Fig. 3.**
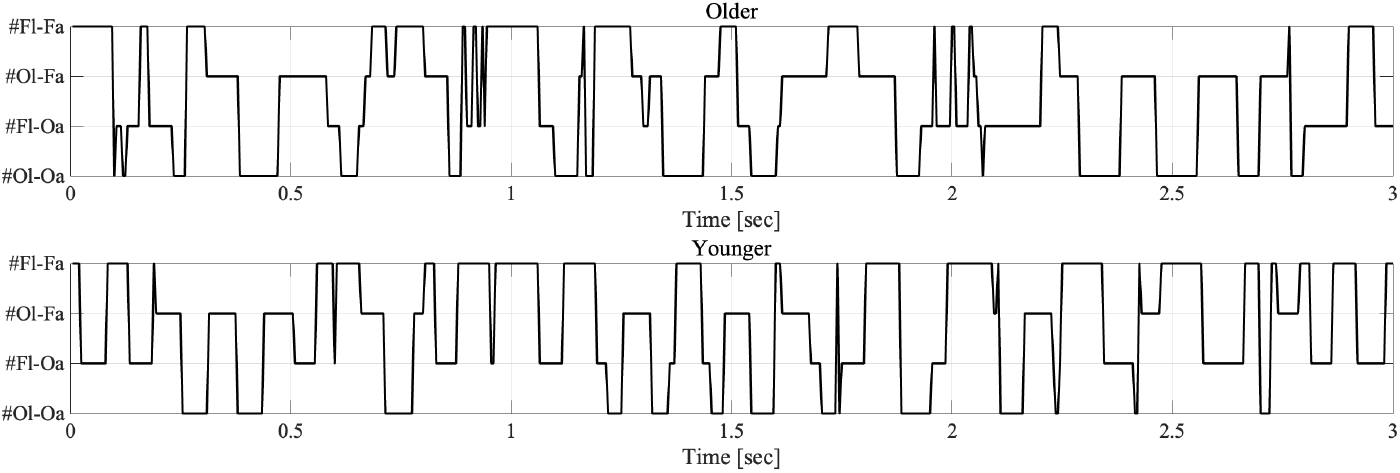
Time series of state transitions among #Ol-Oa, #Fl-Oa, #Ol-Fa, and #Fl-Fa for an older participant (top) and a younger participant (bottom).

**Fig. 4.**
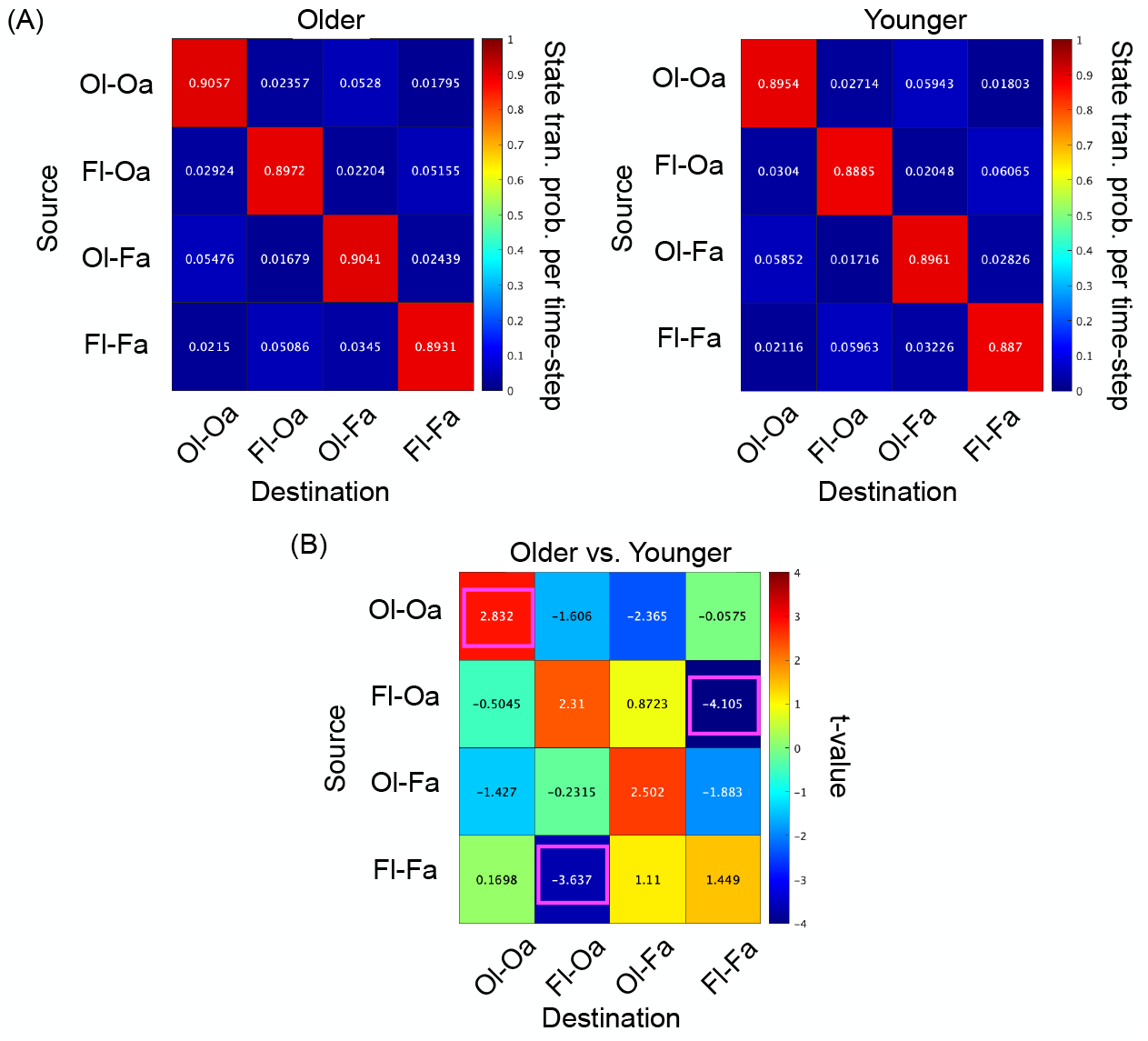
(A) Mean state transition probabilities in the younger and older groups. (B) *t*-values representing group differences in transition probabilities. Positive *t*-values indicate higher probabilities in the older group compared to the younger group; negative values indicate the opposite. Magenta squares indicate transitions with statistically significant differences after Benjamini-Hochberg false discovery rate (FDR) correction (*q <* 0.05). In the older group, transitions from #Fl-Fa to #Fl-Oa and from #Fl-Oa to #Fl-Fa were significantly less frequent; however, the probability of remaining in state #Ol-Oa was significantly greater.

**Fig. 5.**
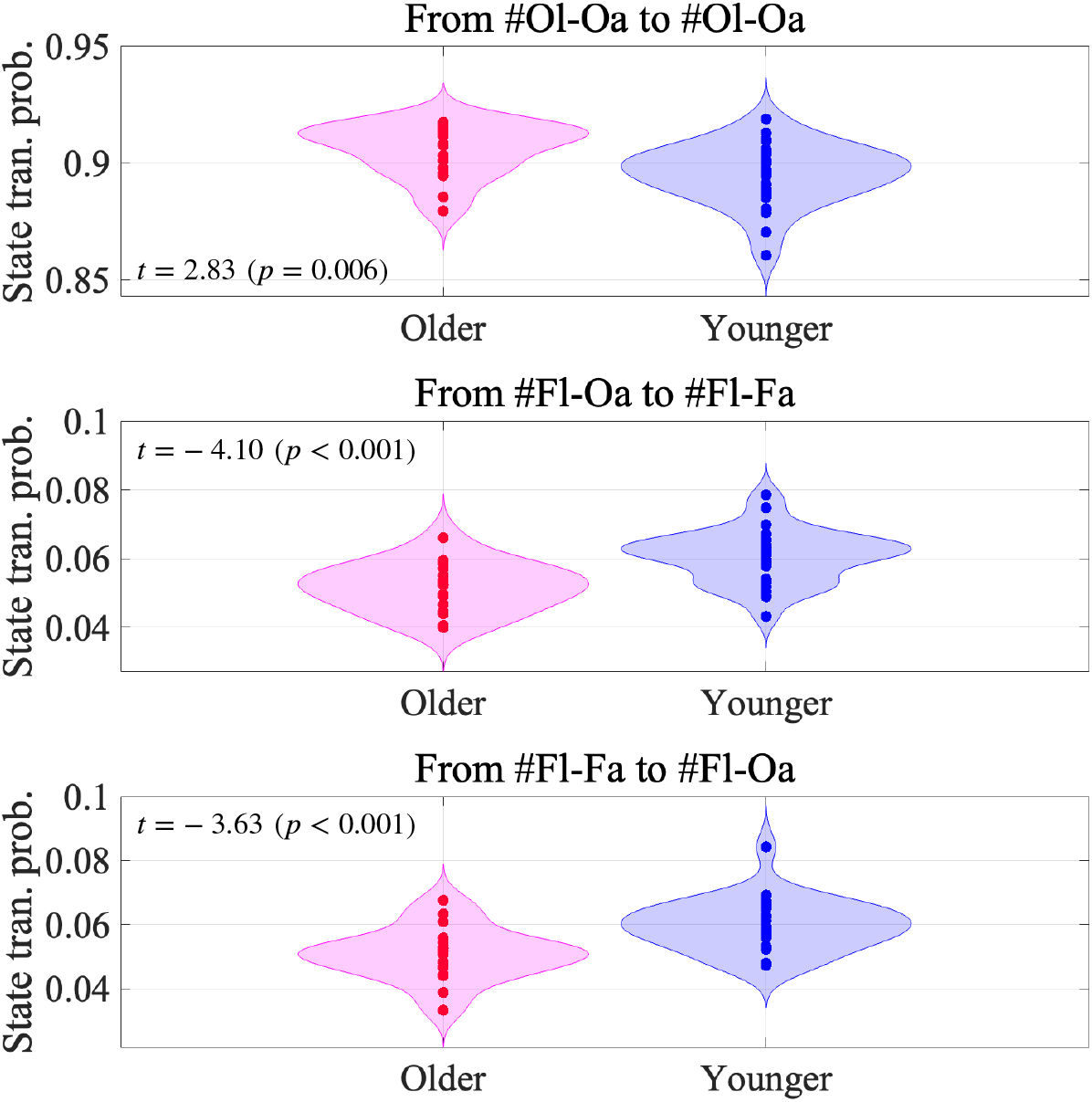
Scatter plots illustrating state transition probabilities from #Fl-Fa to #Fl-Oa and from #Fl-Oa to #Fl-Fa, and the probability of remaining in state #Ol-Oa for younger (blue points) and older (red points) participants. These state transitions exhibit significant differences (*q <* 0.05) between younger and older groups, as shown in Fig. 4. Violin plots illustrate the distributions of these probabilities within each group.

## 4. Discussion

In this study, we developed a novel microstate framework based on the time series data of IF and IA derived from EEG signals. Using this approach, we identified four distinct spatial patterns of IF and IA distributions across microstates: #Ol-Oa, #Fl-Oa, #Ol-Fa, and #Fl-Fa. We then analyzed the temporal transitions between these microstates and identified aging-specific characteristics. Specifically, transitions from #Fl-Fa to #Fl-Oa and from #Fl-Oa to #Fl-Fa were significantly less frequent in the older group, and the probability of remaining in state #Ol-Oa was significantly higher. Importantly, these differences were not statistically significant when analyzing either IF or IA alone, suggesting that the integration of both features is essential for capturing age-related alterations in brain state dynamics.

Previous studies on aging-related alterations in microstates, such as the work by Jabès *et al*., have reported significant differences in the occurrence frequency and transition patterns of EEG microstates associated with brain regions closely linked to cognitive functions, including frontal and parietal areas [14]. In particular, the frequency of transitions into these specific microstates decreases significantly in older adults [14]. In the present study, we observed that the microstates exhibiting significantly reduced transition probabilities in the older group were all frontal-leading. These results are consistent with previous findings on age-related changes in EEG microstates.

In general, a higher instantaneous frequency (phase-leading) observed in one brain region than in other regions is presumed to reflect information transmission among brain regions [26]. In particular, frontal-leading states likely represent the neural activity involved in top-down control originating from the frontal cortex [19]. Thus, the observed decrease in transitions associated with these frontal-leading states suggests an age-related decline in the precision of top-down regulation. This may help explain why microstates characterized by frontal-leading patterns change with aging. Furthermore, the frequency band analyzed in our study (4-13 Hz) included the theta frequency range. Frontal theta-band oscillations play crucial roles in various cognitive functions [33]. Our findings align well with existing evidence regarding the functional relevance of frontal theta activity and its decline with aging.

We found that the frontal-leading microstates related to aging effects could be further subdivided into frontal-amplified and occipital-amplified states; this division enabled the effective detection of aging effects. Typically, the amplitudes of neural oscillations reflect local information processing within specific brain regions, whereas the phase relationships among regions reflect functional coordination across larger neural networks [31, 37]. Thus, the age-related alterations observed suggest a simultaneous decline in both local neural processing (reflected by amplitude) and global neural coordination (reflected by phase synchronization). Crucially, the integration and simultaneous assessment of the amplitude and phase characteristics achieved by our newly introduced IF-IA microstate approach enable the clear identification of these subtle yet meaningful neural dynamics. These results highlight the practical value of the proposed microstate framework.

Finally, this study had certain limitations. First, we set the number of clusters to *k* = 4 to estimate the IF-IA microstates based on the instantaneous spatial distributions of IF and IA. However, the optimal number of clusters for accurately characterizing IF-IA microstates remains unclear and requires further validation. Second, alterations in network dynamics have been reported in various psychiatric disorders [6, 10]. Therefore, disorder-specific alterations in the dynamic states characterized by the IF-IA microstate approach may also exist and should be explored further. We plan to address these issues in future studies.

## 5. Conclusions

In this study, we proposed a novel EEG microstate approach that integrates IF and IA dynamics. Using this method, we identified aging-specific alterations in microstate transitions. Crucially, this age-related effect emerged by jointly analyzing both the amplitude and frequency components, underscoring the importance of integrating multiple neural features. Our findings offer a promising new perspective for understanding brain network dynamics during healthy aging and provide potential applications for future clinical investigations of age-related disorders.

## Acknowledgment

This work was supported by JSPS KAKENHI, Grant-in-Aid for Scientific Research (B) (Grant Number JP25K03198, SN), Scientific Research (C) (Grant Numbers JP23K06983, TT; JP22K12183, SN), and Transformative Research Areas (A) (Grant Number JP25H02626, SN).

## Data Availability

The datasets generated for this study will not be made publicly available, because informed patient consent will not include a declaration regarding the public availability of the clinical data.

## Competing Interests

The authors declare that the research was conducted in the absence of any commercial or financial relationships that could be construed as a potential conflict of interest.

## Author Contributions

SN, TI, and TT designed the study. SN analyzed the results, wrote the main manuscript text, and prepared the figures. MK conducted the experiments. All the authors reviewed the manuscript.

## Notes

### Competing Interest Statement

The authors have declared no competing interest.

### Author Declarations

All methods were conducted in accordance with the Declaration of Helsinki, and the study protocol was approved by the Ethics Committee of Kanazawa University.

